# Aldosterone-targeted Therapy after Primary Aldosteronism Testing in Resistant Hypertension: A Nationwide Cohort Study

**DOI:** 10.64898/2026.05.16.26353384

**Authors:** Cheng-Hsuan Tsai, Yu-Ching Chang, Chin Chen Chang, Yi-Yao Chang, Uei-Lin Chen, Jeff Shih-Chieh Chueh, Jenifer M Brown, Vin-Cent Wu, Yen-Hung Lin, Anand Vaidya

## Abstract

**Background:** Primary aldosteronism (PA) testing is recommended for patients with resistant hypertension but remains underused, and evidence linking aldosterone-targeted therapy to improved cardiovascular and renal outcomes is limited.

**Methods:** In a nationwide cohort of patients with resistant hypertension between 2001 and 2022, we assessed PA testing and subsequent mineralocorticoid receptor antagonist (MRA) use and adrenalectomy. Among tested patients, time-dependent Cox models were used to assess associations between treatment exposure and mortality, major adverse cardiovascular events (MACE) and renal outcomes.

**Results:** Among 254,338 patients, only 2.0% were tested for PA. Tested patients had a higher prevalence of hypokalemia and cardiometabolic comorbidities. In the overall tested population, MRA use was not associated with lower risks of cardiovascular or renal outcomes. However, when testing resulted in an established PA diagnosis, the use of both MRA (hazard ratio [HR] 0.60, 95% CI 0.42–0.86) and adrenalectomy (HR 0.33, 95% CI 0.20–0.54) were associated with a reduced risk of MACE compared with no aldosterone-targeted therapy. Similar results were observed regarding mortality. Adrenalectomy was associated with lower risk of MACE (HR 0.55, 95% CI 0.30–0.99), all-cause mortality (HR 0.52, 95% CI 0.29–0.93) and renal outcomes (HR 0.37, 95% CI 0.17–0.80) compared with MRA in patients with a diagnosis of PA.

**Conclusions:** PA remains markedly underrecognized in resistant hypertension. Among patients with resistant hypertension who did undergo PA testing with establishment of a PA diagnosis, aldosterone-targeted therapy resulted in lower risk of adverse cardiorenal outcomes and death when compared to conventional antihypertensive therapy.

## Introduction

Hypertension is the leading modifiable risk factor for cardiovascular morbidity and mortality worldwide^1,2^. Among hypertensive patients, resistant hypertension is associated with a markedly increased risk of stroke, myocardial infarction, heart failure, and chronic kidney disease^3,4^. Despite the availability of multiple pharmacological therapies, outcomes in this population remain suboptimal, underscoring the need for mechanism-based approaches to diagnosis and treatment.

Primary aldosteronism (PA) is increasingly recognized as a common and potentially treatable cause of hypertension, with an estimated prevalence of up to 25% or higher in patients with resistant hypertension^5–7^. Excess aldosterone promotes sodium retention, volume expansion, and adverse cardiovascular and renal dysfunction, independent of blood pressure levels^8,9^. Importantly, PA is associated with a higher risk of cardiovascular events compared with essential hypertension, even after adjusting for blood pressure and conventional risk factors^10^. Given its high prevalence and the availability of effective aldosterone-targeted therapies, including mineralocorticoid receptor antagonists (MRA) and adrenalectomy, international guidelines have long recommended testing for PA in patients with resistant hypertension^11,12^.

However, despite these recommendations, real-world implementation of PA testing remains limited worldwide. Prior studies have demonstrated substantial underutilization of testing, even among patients with overt clinical features such as hypokalemia or adrenal incidentaloma^13–19^. As a result, a large proportion of patients with PA likely remain undiagnosed and untreated. More importantly, although targeted therapies have been associated with improved outcomes in patients with confirmed PA, it remains unclear whether testing for PA in patients with resistant hypertension results in improved cardiorenal outcomes or survival. To date, only one study in a U.S. veteran population with treatment-resistant hypertension has evaluated the impact of PA testing, demonstrating increased MRA use and modest reductions in blood pressure^14^. Studies evaluating whether PA testing in resistant hypertension is associated with improved long-term cardiovascular and renal outcomes may help reduce clinician inertia to test and treat this population with aldosterone-targeted therapy.

To address these clinical gaps, we conducted a nationwide cohort study using Taiwan’s National Health Insurance Database. We aimed to (1) evaluate the real-world utilization of PA testing in patients with resistant hypertension, (2) examine its association with subsequent treatment, including MRA use and adrenalectomy, and (3) assess whether exposure to these targeted therapies, modeled as time-varying treatments, was associated with improved long-term clinical outcomes. In addition, we investigated whether the effectiveness of MRA differed according to confirmed PA status, as a proxy for biologically relevant aldosterone excess, to clarify the role of diagnostic evaluation in guiding precision therapy in resistant hypertension.

## Methods

### Data source

This nationwide, population-based retrospective cohort study was conducted using the Taiwan National Health Insurance Research Database (NHIRD). The National Health Insurance (NHI) program is a single-payer system with mandatory enrollment, covering more than 99.9% of Taiwan’s population. The NHIRD contains de-identified, longitudinal healthcare data, including diagnostic codes, procedures, and prescription records for all insured individuals. For this study, data were accessed through the Health and Welfare Data Science Center, which enables linkage to external datasets, including the National Death Registry, allowing comprehensive outcome ascertainment. Additional details regarding the NHIRD have been described previously^20–22^. This study was approved by the Institutional Review Board of National Taiwan University Hospital, with a waiver of informed consent due to the use of de-identified data (202508048W).

### Study population

We obtained access to NHIRD data from 2000 to 2023. Diagnostic coding was based on the International Classification of Diseases, Ninth Revision, Clinical Modification (ICD-9-CM) before 2016 and the Tenth Revision (ICD-10-CM) thereafter. Patients with incident resistant hypertension were included from 2001 to 2022. A 12-month look-back period was utilized for baseline covariate identification, while a one-year landmark was set to monitor subsequent outcomes. To ensure inclusion of incident cases, individuals with a diagnosis of hypertension in 2000 were not included. Prior validation studies have demonstrated high accuracy of hypertension identification in the NHIRD, with an accuracy of 93% using diagnosis-based definitions and 95% using medication-based definitions^23^. Resistant hypertension was defined as a diagnosis of hypertension in combination with the concurrent use of at least four classes of antihypertensive medications for one consecutive year^14,24^.

PA testing was defined by the concurrent measurement of plasma renin (either plasma renin activity or direct renin concentration) and plasma aldosterone concentration on the same day, identified using Taiwan NHI reimbursement codes. Patients were excluded based on the following criteria: missing information regarding the institution of hypertension diagnosis, age under 18 years, or a history of end-stage renal disease (ESRD) requiring chronic dialysis. To ensure the inclusion of a newly diagnosed population, we further excluded individuals with prior PA testing or diagnosis, as well as those who had previously received MRA therapy or adrenalectomy before the index date. Exposure to MRA and adrenalectomy history were ascertained using records available from January 1, 2000 onward.

### Exposure to MRA and adrenalectomy

The primary objective of this study was to examine, among patients with resistant hypertension who underwent PA testing, the associations of subsequent MRA therapy and adrenalectomy with subsequent clinical events. Because treatments occurred after PA testing, exposure classification based on future treatment status could introduce immortal time bias. Therefore, we applied a time-dependent analytic approach to mitigate immortal time bias (the Mantel–Byar method)^25^. Each patient’s follow-up could be partitioned into up to three consecutive intervals, for example: from PA testing to MRA initiation, from MRA initiation to adrenalectomy, and from adrenalectomy to the end of follow-up. Because adrenalectomy represents a more definitive, irreversible treatment, patients who underwent adrenalectomy were classified as adrenalectomy-exposed thereafter; any subsequent MRA prescriptions after adrenalectomy were not further considered. Furthermore, to investigate whether the therapeutic benefits of aldosterone-targeted therapy were more pronounced in patients with a more definitive PA pathophysiology, we performed subgroup analyses restricted to those who received a concurrent diagnosis of PA diagnostic code at the time of MRA initiation or adrenalectomy.

### Covariates

Comorbidities were defined using standard claims-based algorithms. Hypokalemia was defined as ≥2 outpatient diagnoses or 1 inpatient diagnosis prior to the index date. Chronic kidney disease and obstructive sleep apnea were defined as ≥2 outpatient diagnoses or 1 inpatient diagnosis within 1 year before the index date. Stroke was defined as any prior hospitalization with a corresponding diagnosis. Additional covariates included age, sex, monthly income (categorized into tertiles), urbanization level of residence and comorbid conditions, including diabetes mellitus, hyperlipidemia, atrial fibrillation, heart failure hospitalization, prior acute myocardial infarction, coronary artery disease, peripheral vascular disease, and chronic obstructive pulmonary disease. We also captured the classes of antihypertensive medications and the total number of antihypertensive agents. Notably, all of the covariates were updated at PA testing, MRA initiation, and adrenalectomy, and were therefore treated as time-varying covariates. Detailed ICD codes are provided in **Supplementary Table S1**.

### Outcomes

The primary outcomes were all-cause mortality, cardiovascular mortality, major adverse cardiovascular events (MACE), and composite renal outcomes. MACE was defined as a composite of cardiovascular death, acute myocardial infarction, and ischemic stroke, with each component also analyzed separately. Additional cardiovascular outcomes included heart failure hospitalization and hemorrhagic stroke.

Composite renal outcomes included advanced chronic kidney disease (CKD) and end-stage renal disease (ESRD) requiring dialysis. All-cause and cardiovascular mortality were ascertained through linkage with the National Death Registry. Study outcomes were defined by principal discharge diagnoses of acute myocardial infarction, ischemic stroke, hospitalization for heart failure, and hemorrhagic stroke.

Advanced CKD was identified by a CKD diagnosis plus the prescription of erythropoiesis-stimulating agents, a proxy for estimated glomerular filtration rate <15 mL/min/1.73 m^2^ based on Taiwan’s NHI reimbursement regulations^26^. ESRD was defined as the commencement of maintenance dialysis, validated through Catastrophic Illness Certificate Database. Patients were followed until the occurrence of the outcome of interest, death, or the end of data availability (December 31, 2023), whichever occurred first.

### Statistical analysis

Baseline characteristics of patients with and without PA testing were compared using the chi-square test for categorical variables and the independent samples t-test for continuous variables. The primary analyses comprised two parts. First, we evaluated the association between post–PA testing MRA use and outcomes regardless of PA status. Second, we examined the associations of post–PA testing MRA use and adrenalectomy with outcomes, restricted to treatment episodes in which a PA diagnosis was recorded at the time of therapy. Exposure to MRA and adrenalectomy was modeled as time-varying covariates, such that patients contributed person-time to the non-exposed group until treatment initiation. Multivariable Cox proportional hazards models were used to estimate hazard ratios (HR) and 95% confidence intervals (CI) for all outcomes, adjusting for age, sex, income level, urbanization, comorbidities and antihypertensive medication burden updated at each exposure time point. All analyses were conducted using SAS version 9.4 (SAS Institute Inc., Cary, NC, USA), and a two-sided *P* value <0.05 was considered statistically significant.

## Results

### Study population and PA testing utilization

A total of 254,338 patients with resistant hypertension were included. Among them, only 5,113 patients (2.0%) underwent PA testing. Compared with those without testing, tested patients had a markedly higher prevalence of hypokalemia (14.6% vs. 1.1%) and a greater burden of cardiometabolic comorbidities, including diabetes mellitus (57.3% vs. 45.3%), coronary artery disease (36.8% vs. 28.9%), and chronic kidney disease (43.7% vs. 23.5%). These patients also received a higher number of antihypertensive medications at baseline (4.3 vs. 4.1), indicating more severe hypertensive disease (**Table 1**).

**Table 1.** Baseline demographic and clinical characteristics of resistant hypertension patients with and without PA testing.

| Variable | Total<br>(*n* = 254,338) | PA testing<br>(*n* = 5,113) | Non-PA testing<br>(*n* = 249,225) | *P* value |
| --- | --- | --- | --- | --- |
| Age, year | 67.4 ± 12.3 | 66.8 ± 12.9 | 67.4 ± 12.3 | 0.001 |
| Male | 165,724 (65.2) | 3,272 (64.0) | 162,452 (65.2) | 0.077 |
| Urbanization level of the residence |  |  |  | <0.001 |
| Low | 30,118 (11.8) | 461 (9.0) | 29,657 (11.9) |  |
| Moderate | 80,317 (31.6) | 1,394 (27.3) | 78,923 (31.7) |  |
| High | 74,521 (29.3) | 1,542 (30.2) | 72,979 (29.3) |  |
| Very high | 69,382 (27.3) | 1,716 (33.6) | 67,666 (27.2) |  |
| Income level |  |  |  | <0.001 |
| Tertile 1 | 85,059 (33.4) | 1,674 (32.7) | 83,385 (33.5) |  |
| Tertile 2 | 85,999 (33.8) | 1,612 (31.5) | 84,387 (33.9) |  |
| Tertile 3 | 83,280 (32.7) | 1,827 (35.7) | 81,453 (32.7) |  |
| Hypokalemia at baseline | 3,422 (1.4) | 747 (14.6) | 2,675 (1.1) |  |
| Comorbidities at diagnosis of resistant hypertension |  |  |  |  |
| Diabetes mellitus | 115,708 (45.5) | 2,928 (57.3) | 112,780 (45.3) | <0.001 |
| Hyperlipidemia | 116,477 (45.8) | 2,805 (54.9) | 113,672 (45.6) | <0.001 |
| Atrial fibrillation | 8,601 (3.4) | 304 (6.0) | 8,297 (3.3) | <0.001 |
| Heart failure | 14,094 (5.5) | 466 (9.1) | 13,628 (5.5) | <0.001 |
| Acute myocardial infarction | 8,627 (3.4) | 235 (4.6) | 8,392 (3.4) | <0.001 |
| Coronary artery disease | 73,950 (29.1) | 1,880 (36.8) | 72,070 (28.9) | <0.001 |
| Peripheral vascular disease | 10,607 (4.2) | 324 (6.3) | 10,283 (4.1) | <0.001 |
| Intracranial hemorrhage | 10,390 (4.1) | 224 (4.4) | 10,166 (4.1) | 0.280 |
| Ischemic stroke | 30,911 (12.2) | 797 (15.6) | 30,114 (12.1) | <0.001 |
| Obstructive sleep apnea | 1,983 (0.8) | 113 (2.2) | 1,870 (0.8) | <0.001 |
| Chronic obstructive pulmonary disease | 18,234 (7.2) | 439 (8.6) | 17,795 (7.1) | <0.001 |
| Chronic kidney disease | 60,686 (23.9) | 2,235 (43.7) | 58,451 (23.5) | <0.001 |
| Antihypertensive medications at baseline |  |  |  |  |
| RASI | 237,616 (93.4) | 4,656 (91.1) | 232,960 (93.5) | <0.001 |
| Beta-blocker | 210,822 (82.9) | 4,272 (83.6) | 206,550 (82.9) | 0.205 |
| Alpha-blocker | 112,010 (44.0) | 2,480 (48.5) | 109,530 (44.0) | <0.001 |
| Vasodilator (hydralazine, minoxidil) | 8,437 (3.3) | 409 (8.0) | 8,028 (3.2) | <0.001 |
| Thiazide | 167,084 (65.7) | 2,929 (57.3) | 164,155 (65.9) | <0.001 |
| Calcium channel blocker | 236,309 (92.9) | 4,695 (91.8) | 231,614 (92.9) | 0.002 |
| Others (clonidine, minoxidil) | 3,231 (1.3) | 210 (4.1) | 3,021 (1.2) | <0.001 |
| Number of antihypertensive medications | 4.1 ± 0.3 | 4.3 ± 0.4 | 4.1 ± 0.3 | <0.001 |
| Follow up duration, years | 6.9 ± 4.9 | 5.8 ± 4.5 | 6.9 ± 4.9 | <0.001 |

Abbreviation: PA, primary aldosteronism; RASI, renin-angiotensin system inhibitors.
Data are presented as frequency (percentage) or mean ± standard deviation.

### Treatment patterns after PA testing

Among patients who underwent PA testing, 2,063 (40.3%) received MRA therapy, whereas only 273 (5.3%) underwent adrenalectomy. The median intervals from PA testing to subsequent MRA initiation and adrenalectomy were 0.6 years (interquartile range [IQR], 0.1–2.5) and 0.4 years (IQR, 0.1–1.2), respectively. Patients undergoing adrenalectomy were substantially younger (58.6 vs. 68.8 years for MRA) and had a higher prevalence of hypokalemia (46.2% vs. 23.6%) (**Table 2**). In contrast, among patients who did not undergo PA testing, 54,885 (22.0%) received MRA therapy and only 73 (<0.01%) underwent adrenalectomy. Although all patients met criteria for resistant hypertension, aldosterone-targeted therapy was less frequently used in those without PA testing than in those who underwent PA testing. Notably, the majority of tested patients still did not receive any aldosterone-targeted therapy.

**Table 2.** Demographic and clinical characteristics of PA-tested patients at PA testing, MRA initiation, and adrenalectomy.

| Variable | At PA testing<br>(*n* = 5,113) | At MRA prescription<br>(*n* = 2,063) | At adrenalectomy<br>(*n* = 273) |
| --- | --- | --- | --- |
| Age, year | 66.8 ± 12.9 | 68.8 ± 12.5 | 58.6 ± 10.9 |
| Male | 3,272 (64.0) | 1,296 (62.8) | 182 (66.7) |
| Urbanization level of the residence |  |  |  |
| Low | 461 (9.0) | 184 (8.9) | 22 (8.1) |
| Moderate | 1,394 (27.3) | 561 (27.2) | 79 (28.9) |
| High | 1,542 (30.2) | 618 (30.0) | 85 (31.1) |
| Very high | 1,716 (33.6) | 700 (33.9) | 87 (31.9) |
| Income level |  |  |  |
| Tertile 1 | 1,699 (33.2) | 703 (34.1) | 55 (20.2) |
| Tertile 2 | 1,755 (34.3) | 688 (33.4) | 99 (36.3) |
| Tertile 3 | 1,658 (32.4) | 672 (32.6) | 119 (43.6) |
| Hypokalemia at baseline | 747 (14.6) | 486 (23.6) | 126 (46.2) |
| Comorbidities at diagnosis of resistant hypertension |  |  |  |
| Diabetes mellitus | 2,928 (57.3) | 1,225 (59.4) | 137 (50.2) |
| Hyperlipidemia | 2,805 (54.9) | 1,099 (53.3) | 141 (51.7) |
| Atrial fibrillation | 304 (6.0) | 194 (9.4) | 11 (4.0) |
| Heart failure | 466 (9.1) | 430 (20.8) | 11 (4.0) |
| Acute myocardial infarction | 235 (4.6) | 145 (7.0) | 4 (1.5) |
| Coronary artery disease | 1,880 (36.8) | 816 (39.6) | 81 (29.7) |
| Peripheral vascular disease | 324 (6.3) | 158 (7.7) | 12 (4.4) |
| Intracranial hemorrhage | 224 (4.4) | 134 (6.5) | 14 (5.1) |
| Ischemic stroke | 797 (15.6) | 367 (17.8) | 15 (5.5) |
| Obstructive sleep apnea | 113 (2.2) | 67 (3.3) | 5 (1.8) |
| Chronic obstructive pulmonary disease | 439 (8.6) | 221 (10.7) | 12 (4.4) |
| Chronic kidney disease | 2,235 (43.7) | 973 (47.2) | 98 (35.9) |
| Antihypertensive medications |  |  |  |
| RASI | 4,656 (91.1) | 1,565 (75.9) | 219 (80.2) |
| Beta-blocker | 4,272 (83.6) | 1,421 (68.9) | 206 (75.5) |
| Alpha-blocker | 2,480 (48.5) | 878 (42.6) | 106 (38.8) |
| Vasodilator (hydralazine, minoxidil) | 409 (8.0) | 157 (7.6) | 14 (5.1) |
| Thiazide | 2,929 (57.3) | 791 (38.3) | 113 (41.4) |
| Calcium channel blocker | 4,695 (91.8) | 1,612 (78.1) | 229 (83.9) |
| Others (clonidine, minoxidil) | 210 (4.1) | 83 (4.0) | 7 (2.6) |
| Number of antihypertensive medications | 4.3 ± 0.4 | 4.0 ± 1.0 | 4.0 ± 0.8 |
| Follow up duration, years | 3.5 ± 3.8 | 4.7 ± 4.2 | 7.5 ± 5.2 |

Abbreviation: PA, primary aldosteronism; RASI, renin-angiotensin system inhibitors;
Data are presented as frequency (percentage) or mean ± standard deviation.

### Association of MRA with outcomes

In time-dependent analyses among all tested patients, MRA use was not associated with improved clinical outcomes compared with no aldosterone-targeted therapy (**Figure 2A** and **Table 3**). Specifically, MRA use was not associated with lower risks of all-cause mortality (hazard ratio [HR], 0.95; 95% confidence interval [CI], 0.86–1.06), cardiovascular mortality (HR, 0.98; 95% CI, 0.85–1.13), or MACE (HR, 0.99; 95% CI, 0.88–1.12). However, MRA use was associated with a lower risk of hemorrhagic stroke (HR, 0.69; 95% CI, 0.53–0.91).

**Figure 1.**
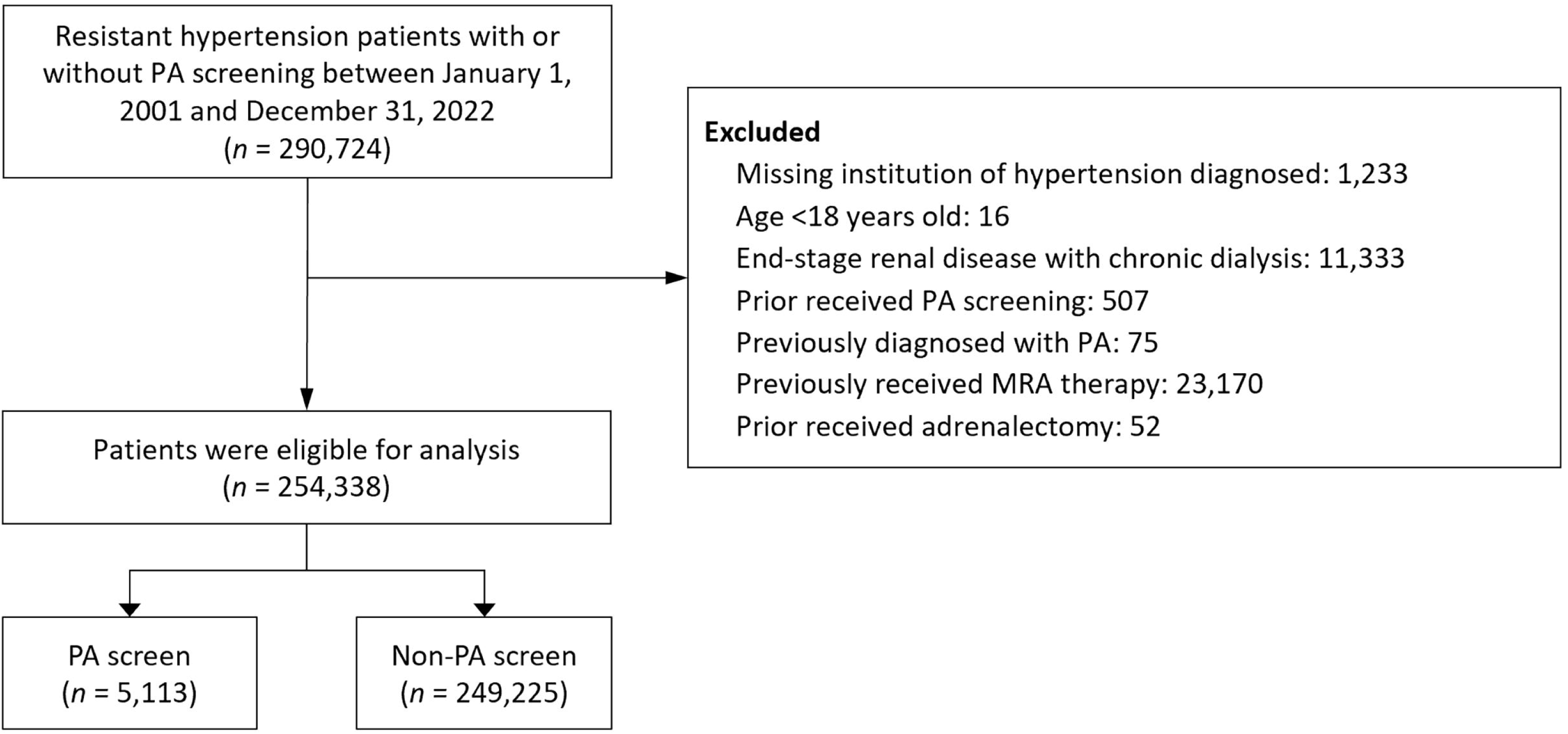
Flowchart of study participant selection, including inclusion and exclusion criteria. PA, primary aldosteronism.

**Figure 2.**
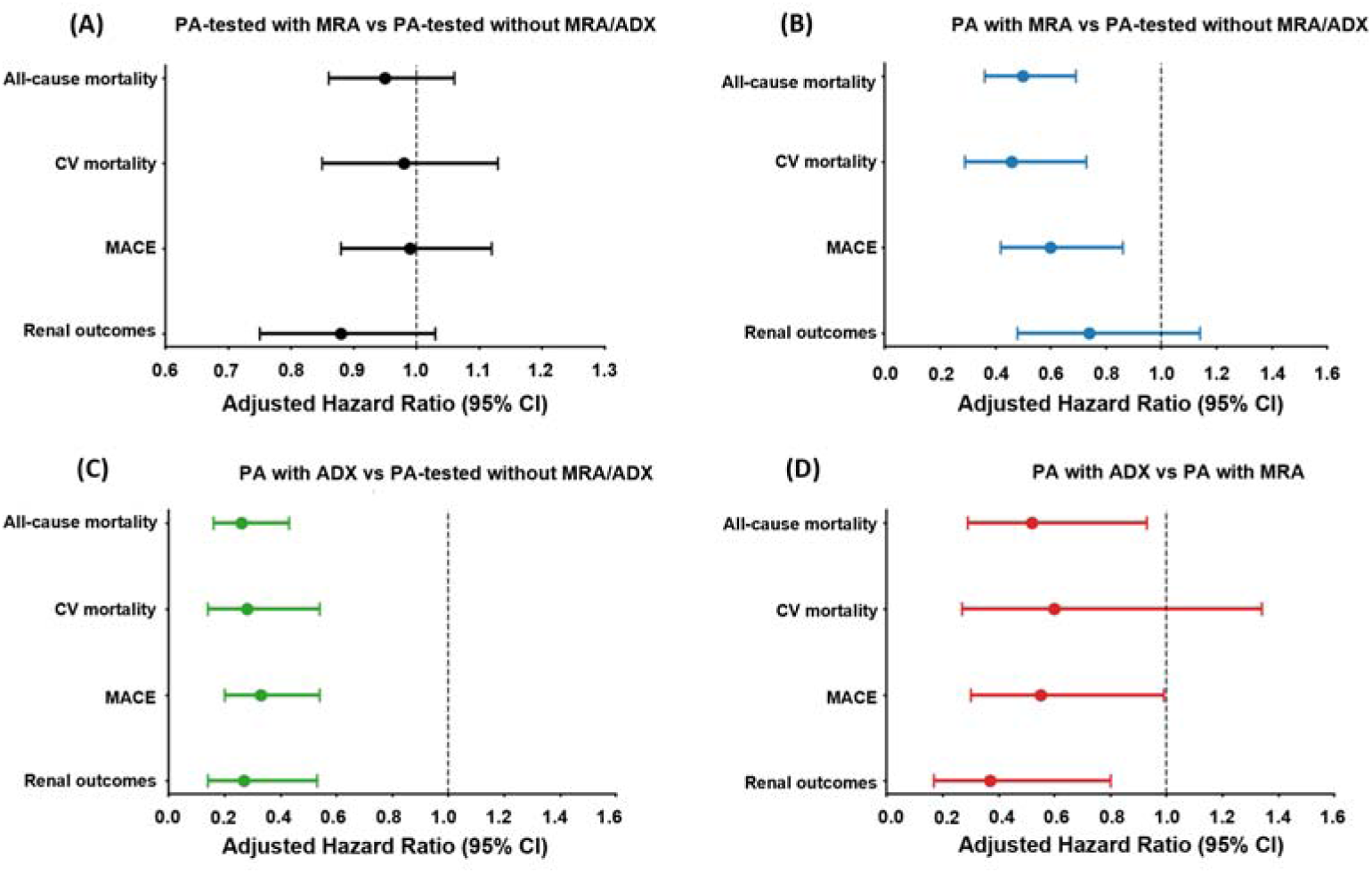
Association of aldosterone-targeted therapies with clinical outcomes in patients with resistant hypertension undergoing PA testing. (A) Adjusted hazard ratios (HRs) for clinical outcomes associated with time-varying MRA use compared with no aldosterone-targeted treatment in the overall PA-tested population. (B) Adjusted HRs for clinical outcomes associated with time-varying MRA use among patients with an established PA diagnosis, using patients without aldosterone-targeted treatment in the overall PA-tested population as the reference group. (C) Adjusted HRs for clinical outcomes associated with adrenalectomy among patients with PA, using patients without aldosterone-targeted treatment in the overall PA-tested population as the reference group. (D) Adjusted HRs comparing adrenalectomy versus MRA among patients with PA. **Abbreviation:** ADX, adrenalectomy; MRA, mineralocorticoid receptor antagonist; PA, primary aldosteronism.

**Table 3.** Association between time-varying MRA exposure and the risk of clinical outcomes.

| Outcome | Incidence (95% CI)<sup>a</sup> |  | Univariate analysis |  | Multivariable analysis<sup>b</sup> |  |
| --- | --- | --- | --- | --- | --- | --- |
|  | Non-MRA | MRA | HR (95% CI) | *P* value | HR (95% CI) | *P* value |
| All-cause mortality | 6.0 (5.6–6.3) | 8.2 (7.6–8.7) | 1.37 (1.25–1.50) | <0.001 | 0.95 (0.86–1.06) | 0.378 |
| Cardiovascular mortality | 3.0 (2.7–3.3) | 4.4 (4.0–4.9) | 1.47 (1.30–1.67) | <0.001 | 0.98 (0.85–1.13) | 0.777 |
| MACE<sup>c</sup> | 5.5 (5.1–5.8) | 6.8 (6.3–7.4) | 1.25 (1.13–1.39) | <0.001 | 0.99 (0.88–1.12) | 0.906 |
| MACCE<sup>d</sup> | 6.0 (5.6–6.3) | 7.0 (6.5–7.6) | 1.19 (1.07–1.31) | 0.001 | 0.95 (0.85–1.07) | 0.370 |
| Acute myocardial infarction | 1.2 (1.1–1.4) | 1.5 (1.3–1.8) | 1.26 (1.02–1.56) | 0.034 | 1.00 (0.79–1.28) | 0.985 |
| Ischemic stroke | 2.1 (1.9–2.3) | 2.4 (2.1–2.8) | 1.20 (1.01–1.42) | 0.037 | 1.01 (0.84–1.23) | 0.906 |
| Hemorrhagic stroke | 1.2 (1.0–1.3) | 1.0 (0.8–1.3) | 0.88 (0.69–1.12) | 0.290 | 0.69 (0.53–0.91) | 0.008 |
| Heart failure hospitalization | 4.3 (4.0–4.6) | 5.3 (4.7–5.8) | 1.22 (1.08–1.37) | 0.002 | 1.10 (0.96–1.26) | 0.170 |
| Advanced CKD | 3.6 (3.3–3.9) | 3.2 (2.8–3.6) | 0.90 (0.78–1.04) | 0.147 | 0.88 (0.74–1.03) | 0.110 |
| ESRD with chronic dialysis | 2.8 (2.5–3.0) | 2.7 (2.4–3.0) | 0.99 (0.84–1.16) | 0.876 | 0.91 (0.76–1.09) | 0.292 |
| Composite renal outcome | 3.8 (3.5–4.1) | 3.4 (3.0–3.8) | 0.90 (0.79–1.04) | 0.154 | 0.88 (0.75–1.03) | 0.106 |

Abbreviation: CI, confidence interval; CKD, chronic kidney disease; ESRD, end-stage renal disease; HR, hazard ratio; MACCE, major adverse cardiac and cerebrovascular events; MACE, major adverse cardiovascular events; MRA, mineralocorticoid receptor antagonists.
a Number of events per 100 person-years.
b Adjusted for sex, age, urbanization level of the residence, income level, hypokalemia at baseline, diabetes mellitus, hyperlipidemia, atrial fibrillation, heart failure, acute myocardial infarction, coronary artery disease, peripheral vascular disease, intracranial hemorrhage, ischemic stroke, obstructive sleep apnea, chronic obstructive pulmonary disease, chronic kidney disease, number of antihypertensive medications.
c Any of cardiovascular death, acute myocardial infarction, and ischemic stroke.
d Any of cardiovascular death, acute myocardial infarction, ischemic stroke, and heart failure hospitalization.

### Aldosterone-targeted therapy with established PA diagnosis

During the study period, 383 of 5,113 tested patients (7.5%) had a recorded diagnosis of PA. When analyses were restricted to patients with an established PA diagnosis, both MRA use and adrenalectomy were significantly associated with improved outcomes (**Figure 2B**, **Figure 2C** and **Table 4**). When a PA diagnosis had been established, MRA use was associated with lower risks of all-cause mortality (HR, 0.50; 95% CI, 0.36–0.69), cardiovascular mortality (HR, 0.46; 95% CI, 0.29–0.73), and MACE (HR, 0.60; 95% CI, 0.42–0.86) compared with periods without aldosterone-targeted therapy among PA-tested patients (**Figure 2B**). In addition, compared with PA-tested periods without aldosterone-targeted therapy, MRA use was associated with a greater benefit for advanced CKD (HR 0.62, 95% CI 0.39–0.99), whereas associations with composite renal outcomes and ESRD were not statistically significant. Importantly, compared with MRA, adrenalectomy was associated with lower risks of MACE (HR, 0.55; 95% CI, 0.30–0.99), all-cause mortality (HR, 0.52; 95% CI, 0.29–0.93), and composite renal outcomes (HR, 0.37; 95% CI, 0.17–0.80) in patients with PA (**Figure 2D**). The unadjusted hazard ratios for the comparison of study outcomes are provided in the supplements (**Supplementary Table 2**).

**Table 4.**
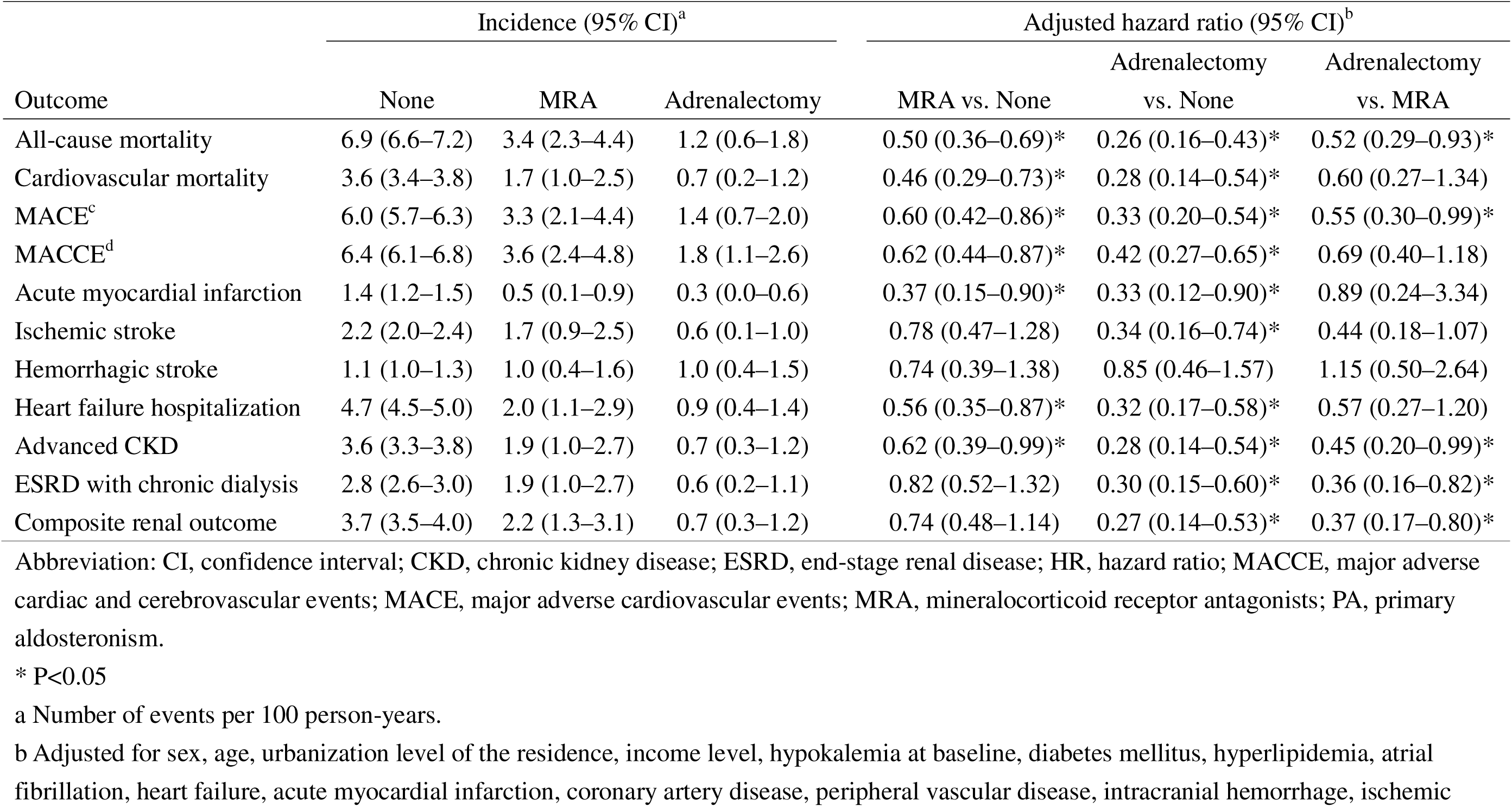
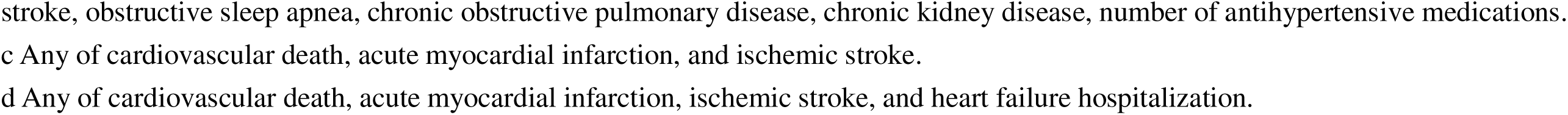
Association of time-varying MRA exposure and adrenalectomy with risks of clinical outcomes, restricted to treatment episodes with a PA diagnosis.

| Outcome | Incidence (95% CI)<sup>a</sup> |  |  | Adjusted hazard ratio (95% CI)<sup>b</sup> |  |  |
| --- | --- | --- | --- | --- | --- | --- |
|  | None | MRA | Adrenalectomy | MRA vs. None | Adrenalectomy vs. None | Adrenalectomy vs. MRA |
| All-cause mortality | 6.9 (6.6–7.2) | 3.4 (2.3–4.4) | 1.2 (0.6–1.8) | 0.50 (0.36–0.69)* | 0.26 (0.16–0.43)* | 0.52 (0.29–0.93)* |
| Cardiovascular mortality | 3.6 (3.4–3.8) | 1.7 (1.0–2.5) | 0.7 (0.2–1.2) | 0.46 (0.29–0.73)* | 0.28 (0.14–0.54)* | 0.60 (0.27–1.34) |
| MACE<sup>c</sup> | 6.0 (5.7–6.3) | 3.3 (2.1–4.4) | 1.4 (0.7–2.0) | 0.60 (0.42–0.86)* | 0.33 (0.20–0.54)* | 0.55 (0.30–0.99)* |
| MACCE<sup>d</sup> | 6.4 (6.1–6.8) | 3.6 (2.4–4.8) | 1.8 (1.1–2.6) | 0.62 (0.44–0.87)* | 0.42 (0.27–0.65)* | 0.69 (0.40–1.18) |
| Acute myocardial infarction | 1.4 (1.2–1.5) | 0.5 (0.1–0.9) | 0.3 (0.0–0.6) | 0.37 (0.15–0.90)* | 0.33 (0.12–0.90)* | 0.89 (0.24–3.34) |
| Ischemic stroke | 2.2 (2.0–2.4) | 1.7 (0.9–2.5) | 0.6 (0.1–1.0) | 0.78 (0.47–1.28) | 0.34 (0.16–0.74)* | 0.44 (0.18–1.07) |
| Hemorrhagic stroke | 1.1 (1.0–1.3) | 1.0 (0.4–1.6) | 1.0 (0.4–1.5) | 0.74 (0.39–1.38) | 0.85 (0.46–1.57) | 1.15 (0.50–2.64) |
| Heart failure hospitalization | 4.7 (4.5–5.0) | 2.0 (1.1–2.9) | 0.9 (0.4–1.4) | 0.56 (0.35–0.87)* | 0.32 (0.17–0.58)* | 0.57 (0.27–1.20) |
| Advanced CKD | 3.6 (3.3–3.8) | 1.9 (1.0–2.7) | 0.7 (0.3–1.2) | 0.62 (0.39–0.99)* | 0.28 (0.14–0.54)* | 0.45 (0.20–0.99)* |
| ESRD with chronic dialysis | 2.8 (2.6–3.0) | 1.9 (1.0–2.7) | 0.6 (0.2–1.1) | 0.82 (0.52–1.32) | 0.30 (0.15–0.60)* | 0.36 (0.16–0.82)* |
| Composite renal outcome | 3.7 (3.5–4.0) | 2.2 (1.3–3.1) | 0.7 (0.3–1.2) | 0.74 (0.48–1.14) | 0.27 (0.14–0.53)* | 0.37 (0.17–0.80)* |

Abbreviation: CI, confidence interval; CKD, chronic kidney disease; ESRD, end-stage renal disease; HR, hazard ratio; MACCE, major adverse cardiac and cerebrovascular events; MACE, major adverse cardiovascular events; MRA, mineralocorticoid receptor antagonists; PA, primary aldosteronism.
\* P<0.05
a Number of events per 100 person-years.
b Adjusted for sex, age, urbanization level of the residence, income level, hypokalemia at baseline, diabetes mellitus, hyperlipidemia, atrial fibrillation, heart failure, acute myocardial infarction, coronary artery disease, peripheral vascular disease, intracranial hemorrhage, ischemic
stroke, obstructive sleep apnea, chronic obstructive pulmonary disease, chronic kidney disease, number of antihypertensive medications.
c Any of cardiovascular death, acute myocardial infarction, and ischemic stroke.
d Any of cardiovascular death, acute myocardial infarction, ischemic stroke, and heart failure hospitalization.

## Discussion

In this nationwide cohort of patients with resistant hypertension, we found that testing for PA remains markedly limited, with only 2.0% of patients undergoing evaluation. We also observed that PA testing was associated with increased use of aldosterone-targeted therapies; however, the effectiveness of these therapies appeared to depend on the establishment of a PA diagnosis. Specifically, while MRA therapy was not associated with improved outcomes in the overall tested population, significant reductions in mortality and cardiovascular risks were observed when treatment was given to those who also received an established PA diagnosis.

Adrenalectomy was consistently associated with the greatest risk reduction across cardiovascular and renal outcomes. In addition, among patients with established PA, adrenalectomy was associated with lower cardiovascular and renal outcomes compared with MRA therapy. These findings underscore the value of PA testing in resistant hypertension by demonstrating its association with increased PA diagnosis and greater use of aldosterone-targeted therapy, which in turn was associated with lower risks of cardiorenal outcomes and death among patients with established PA. These findings are consistent with current guideline recommendations supporting the use of MRA as an effective and pragmatic treatment option, particularly in settings where access to subtype classification and surgical intervention is limited^27–29^.

Our findings extend prior evidence regarding the clinical impact of PA testing. In a large U.S. Veterans cohort, testing for PA was similarly infrequent but was associated with a higher likelihood of initiating MRA therapy and modest improvements in blood pressure control in treatment-resistant hypertension^14^. Consistent with these findings, we also observed that patients who underwent PA testing were more likely to receive aldosterone-targeted therapies, including MRA and adrenalectomy. However, the previous study primarily focused on treatment patterns and blood pressure control and did not evaluate long-term cardiovascular or renal outcomes^14^. In contrast, our study provides important new insights by demonstrating that the clinical benefits of aldosterone-targeted therapy are not uniform across all tested patients but are largely confined to those with an established diagnosis of PA. While prior work suggested that testing may facilitate more frequent use of MRA^14^, our results further clarify that such treatment might translate into meaningful reductions in mortality and cardiovascular events only when applied to the appropriate biological subgroup. These findings suggest that the value of PA testing extends beyond increasing treatment utilization and lies in enabling accurate identification of patients with biologically relevant, renin-independent aldosterone excess who are most likely to benefit from MRA and achieve meaningful improvements in long-term clinical outcomes.

The differential effectiveness of aldosterone-targeted therapy observed in our study can be explained by the underlying pathophysiology of PA. PA is characterized by autonomous, renin-independent aldosterone excess, leading to sodium retention, volume expansion, and mineralocorticoid receptor–mediated cardiometabolic and renal injury^30–37^. These effects promote myocardial fibrosis, vascular remodeling, and renal damage, which contribute to the excess cardiovascular risk observed in patients with PA beyond blood pressure alone^10,38^. In this context, MRAs directly counteract the pathogenic effects of aldosterone and therefore represent a mechanism-based therapy for PA. This concept is supported by the PATHWAY-2 trial, which demonstrated that spironolactone was the most effective add-on therapy for resistant hypertension, particularly in patients with suppressed renin levels, suggesting a volume-expanded, aldosterone-driven phenotype^39,40^. Our findings extend this physiological framework by showing that the benefit of MRA therapy is not universal across all patients with resistant hypertension but is largely confined to those with an established diagnosis of PA. The lack of benefit observed in the overall tested population likely reflects the heterogeneity of hypertension mechanisms, whereas the substantial benefit observed in patients with established PA highlights the importance of aligning treatment with the underlying disease biology.

Another important finding of our study is that, among patients with established PA, cardiovascular and renal outcomes were significantly better in those who underwent adrenalectomy than in those treated with MRA. Nevertheless, MRA therapy was still associated with significant long-term risk reduction in patients with resistant hypertension and confirmed PA. Although adrenalectomy has long been recognized as the first-line therapy for patients with unilateral PA, comprehensive diagnostic evaluation, particularly subtype classification using procedures such as adrenal vein sampling, remains limited in clinical practice^27–29^. In this context, our findings support the use of MRA as a practical and effective initial treatment strategy in patients with resistant hypertension and a phenotype suggestive of PA, such as suppressed renin and an elevated aldosterone-to-renin ratio. However, in patients who are suitable surgical candidates and have access to appropriate diagnostic evaluation, early consideration of adrenalectomy remains the gold standard, given its greater and more definitive risk reduction^27,41,42^.

Another important determinant of MRA therapy efficacy is the degree of renin response following treatment initiation. Hundemer et al. reported that MRA therapy was associated with a higher risk of atrial fibrillation and adverse cardiometabolic outcomes when renin remained suppressed, whereas titration of MRA to achieve renin elevation mitigated these risks^43,44^. Similarly, in a large cohort of patients with unilateral PA, Wu et al. demonstrated that optimized MRA therapy was associated with cardiovascular outcomes comparable to those of surgical adrenalectomy, whereas persistently suppressed renin was associated with a higher risk of adverse events^45^.

These findings are further supported by meta-analytic data showing that unsuppressed renin following medical therapy is associated with reduced risks of cardiovascular events and all-cause mortality, suggesting that renin normalization may serve as a therapeutic target^46,47^. In this context, the comparatively attenuated benefit of MRA relative to adrenalectomy observed in our study may reflect suboptimal renin response in real-world practice. Given that our dataset does not capture longitudinal renin measurements or dose titration, it is likely that a substantial proportion of patients did not achieve adequate renin elevation following MRA initiation.

This study has important clinical implications. First, we demonstrate that despite longstanding guideline recommendations, PA testing remains markedly underutilized, even among high-risk populations such as patients with resistant hypertension. Second, we found that the act of PA testing in resistant hypertension was associated with a substantially higher likelihood of receiving aldosterone-targeted therapies, including MRA and adrenalectomy. Third, patients with an established diagnosis of PA, which was used here as a proxy for renin-independent aldosterone excess, derived substantial benefit from aldosterone-targeted therapy, whereas no significant benefit was observed in the overall tested population. These findings indicate that the effectiveness of treatment is closely linked to the underlying disease mechanism and underscore the importance of aligning therapy with disease pathophysiology. Our results therefore support a more systematic, mechanism-based approach to resistant hypertension, in which diagnostic evaluation facilitates the identification of patients most likely to benefit from aldosterone-targeted therapy. Given the limited availability of subtype classification procedures such as adrenal vein sampling, a PA testing-based strategy followed by MRA therapy may represent a pragmatic approach in many real-world settings.

Several limitations of this study should be acknowledged. First, as a retrospective observational study based on administrative claims data, residual confounding cannot be fully excluded despite multivariable adjustment. Important clinical variables, including blood pressure levels, biochemical parameters, and medication adherence, were not available in the NHIRD. Second, the identification of PA relied on an operational definition based on diagnostic codes and treatment patterns rather than standardized biochemical confirmation. Only 7.5% of patients who underwent PA testing had a recorded PA diagnosis, which is lower than the expected prevalence of

PA among patients with resistant hypertension^5,48^. This discrepancy suggests that PA may have remained underdiagnosed even among tested patients, possibly due to variation in diagnostic criteria, the traditional requirement for multiple confirmatory steps before diagnosis, or undercoding in claims data. Therefore, some patients with biologically relevant aldosterone excess may have been misclassified as not having PA. This misclassification could have diluted the observed treatment effect of MRA in the overall tested population and limited our ability to estimate the full benefit of aldosterone-targeted therapy. In addition, a recorded PA diagnosis may also capture differences in clinical recognition and treatment intensity. Clinicians who coded PA may have been more likely to titrate MRA therapy more aggressively, monitor response more closely, or refer patients for adrenalectomy. Therefore, the stronger associations observed among patients with a PA diagnosis may partly reflect differences in management intensity in addition to the biological effect of aldosterone-targeted therapy. Third, treatment with MRA was not randomly assigned.

Patients who received aldosterone-targeted therapy may have had a higher clinical suspicion of PA or more pronounced biochemical features, introducing potential confounding by indication. As a result, the observed treatment effects may reflect, at least in part, differences in underlying disease severity rather than treatment alone, and the direction of bias cannot be definitively determined. Similarly, patients undergoing adrenalectomy were generally younger and had fewer comorbidities, which may have contributed to their more favorable outcomes. Although time-dependent analyses and multivariable adjustment were applied, residual selection bias cannot be excluded. Fourth, resistant hypertension was defined using a claims-based operational definition, based on a diagnosis of hypertension together with the concurrent use of at least four classes of antihypertensive medications for one consecutive year. Because blood pressure values, medication dose, adherence, and out-of-office blood pressure measurements were unavailable, this definition differs slightly from contemporary guideline-confirmed resistant hypertension. Nevertheless, the sustained requirement for four or more antihypertensive classes over one year likely identifies patients with a clinically meaningful treatment-resistant phenotype and is consistent with prior claims-based studies of apparent treatment-resistant hypertension^13,14^. Fifth, MRA exposure was identified based on prescription claims, and detailed information on dosage, titration, and adherence was not available.

Therefore, we were unable to assess whether renin optimization influenced the observed associations. Finally, this study was conducted in a nationwide Taiwanese population within a single-payer healthcare system, which may limit generalizability to other healthcare settings or ethnic populations. Despite these limitations, the consistency of findings across multiple clinically relevant outcomes and subgroups supports the robustness of our main conclusions.

### Perspectives

In this nationwide cohort of patients with resistant hypertension, testing for PA was markedly underutilized. Nevertheless, aldosterone-targeted therapies conferred substantial clinical benefit in patients with PA. Importantly, the effectiveness of MRA therapy was highly dependent on appropriate patient selection, with significant reductions in cardiovascular risk observed only among patients with an established diagnosis of PA. These findings highlight that the clinical value of PA testing lies in identifying a renin-independent aldosterone excess phenotype in resistant hypertension, in whom subsequent aldosterone-targeted therapy represents an effective mechanism-based approach to improve cardiovascular and renal outcomes. In addition, among patients with established PA, both medical and surgical aldosterone-targeted therapies can improve outcomes when applied to appropriately selected individuals, highlighting the critical role of diagnostic evaluation in guiding treatment decisions. Expanding access to PA testing may therefore represent a practical and scalable strategy to enable precision, mechanism-based care and improve long-term outcomes in resistant hypertension.

## Disclosure

AV reports consulting fees unrelated to the contents of this work from Corcept Therapeutics, Mineralys, HRA Pharma, Moderna, SideraBio, Vertex, AstraZeneca, Adaptyx. JMB reports consulting fees unrelated to the contents of this work from Recordati Rare Diseases, Mineralys and AstraZeneca. All other coauthors have nothing to disclose.

## Data Availability

All data produced in the present study are available upon reasonable request to the authors.

## Acknowledgements

We thank the staff of the Eighth and Second Core Lab in the Department of Medical Research at National Taiwan University Hospital for technical support. We also thank all members of the TAIPAI Study Group (https://doi.org/10.6084/m9.figshare.21669929.v7) for help during the study.

## Novelty and Relevance

### What Is New?

- In a nationwide cohort of patients with resistant hypertension, PA testing was markedly underutilized, with only 2% of patients undergoing evaluation despite guideline recommendations.
- MRA therapy was not associated with improved outcomes in the overall tested population but was significantly associated with reduced mortality and cardiovascular risk when restricted to patients with an established diagnosis of PA.
- Adrenalectomy was associated with greater reductions in cardiovascular and renal outcomes compared with MRA therapy among patients with PA, supporting the superiority of definitive, mechanism-targeted intervention.

### What Is Relevant?

- These findings demonstrate that the effectiveness of aldosterone-targeted therapy is highly dependent on appropriate patient selection and underlying disease biology, rather than treatment alone.
- PA testing plays a critical role not only in diagnosis but also in identifying patients with renin-independent aldosterone excess who are most likely to benefit from aldosterone-targeted therapy.
- Empiric use of MRA without consideration of underlying pathophysiology may dilute its therapeutic benefit in resistant hypertension.

### Clinical Implications?

- Improving access to PA testing and diagnostic evaluation remains an urgent priority to enhance cardiovascular and renal outcomes in patients with resistant hypertension.
- Systematic PA testing in resistant hypertension may enable a transition from empiric treatment escalation to a precision, mechanism-based therapeutic approach.

---

**Supplementary Table 1.** Diagnostic codes for diseases used in this study.

| Disease | ICD-9-CM diagnostic code | ICD-10-CM diagnostic code |
| --- | --- | --- |
| Hypertension | 401.xx-405.xx | I10-I15, N262 |
| Primary aldosteronism | 255.1 | E26.01, E26.02, E26.09, E26.89, E26.9 |
| Hypokalemia | 276.8 | E87.6 |
| Diabetes mellitus | 250.x | E08-E13 |
| Hyperlipidemia | 272.0-272.4 | E77, E780, E781, E782, E783, E784, E785, E786, E881, E753, E755, E882, E756, E789, E7521, E7522, E7524, E7130, E7879, E7881, E7889, E8889, E7870 |
| Atrial fibrillation | 427.31 | I480-I482, I4891 |
| Heart failure | 428.x | I50 |
| Acute myocardial infarction | 410.x | I21-I22 |
| Myocardial infarction | 410.x, 412.x | I21-I22, I25.2 |
| Coronary artery disease | 410.x–414.x | I20-I22, I24, I25 |
| Peripheral vascular disease | 440.0x, 440.2x, 440.3x, 440.8x, 440.9x, 443.x, 444.0x, 444.22, 444.8x, 447.8x, 447.9x | I70, I71, I73, I75, I771, I790, I791, I792, I773, I779, I798, K551, K558, K559, Z958, Z959, I743, I744, I745, I748, I740, I7789 |
| Intracranial hemorrhage | 430.x–432.x | I60-I62 |
| Ischemic stroke | 433.x–437.x | G450, G451, G452, G454, G458, G459, G460, G461, G462, G463, G464, G465, G466, G467, G468, I6300, I63011, I63012, I63019, I6302, I6303, I6309, I6310, I63111, |
|  |  | I63112, I63119, I6312, I6313, I6319, I6320, I63211, I63212, I63219, I6322, I6323, I6329, I6330, I6331, I63311, I63312, I63319, I63321, I63322, I63329, I6333, I63331, I63332, I63339, I6334, I63341, I63342, I63349, I6339, I6340, I6341, I63411, I63412, I63419, I6342, I63421, I63422, I63429, I63431, I63432, I63439, I63441, I63442, I63449, I6349, I6350, I63511, I63512, I63519, I63521, I63522, I63529, I63531, I63532, I63539, I63541, I63542, I63549, I6359, I6359, I636, I638, I639, I650, I651, I6521, I6522, I6523, I6529, I658, I659, I66, I670, I671, I672, I674, I675, I676, I677, I6781, I6782, I6789, I679, I680, I682, I688 |
| Obstructive sleep apnea | 327.23, 780.51, 780.53, 780.57 | G47.30, G47.31, G47.33, G47.37, G47.39 |
| Chronic obstructive pulmonary disease | 491.x, 492.x, 496.x | J41-J44 |
| Chronic kidney disease | 580.x–589.x, 403.x–404.x, 016.0x, 095.4x, 236.9x, 250.4x, 274.1x, 442.1x, 447.3x, 440.1x, 572.4x, 642.1x, 646.2x, 753.1x, 283.11, 403.01, 404.02, 446.21 | A1811, D593, E102, E112, E132, I12, I13, K767, M103, M310, N00, N01, N02, N03, N04, N05, N06, N07, N08, N14, N150, N158, N159, N16, N171, N172, N18, N19, N200, N25, N261, N269, N27, Q61 |

Abbreviation: ICD-9-CM, international classification of diseases, ninth revision, clinical modification; ICD-10-CM, international classification of diseases, tenth revision, clinical modification.

**Supplementary Table 2.** Association of time-varying MRA exposure and adrenalectomy with risks of clinical outcomes, restricted to treatment episodes with a PA diagnosis.

| Outcome | MRA vs. None |  | Adrenalectomy vs. None |  | Adrenalectomy vs. MRA |  |
| --- | --- | --- | --- | --- | --- | --- |
|  | HR (95% CI) | *P* value | HR (95% CI) | *P* value | HR (95% CI) | *P* value |
| All-cause mortality | 0.49 (0.35–0.68) | <0.001 | 0.18 (0.11–0.29) | <0.001 | 0.37 (0.21–0.66) | <0.001 |
| Cardiovascular mortality | 0.49 (0.31–0.76) | 0.002 | 0.20 (0.10–0.38) | <0.001 | 0.40 (0.18–0.89) | 0.024 |
| MACE<sup>a</sup> | 0.54 (0.38–0.77) | <0.001 | 0.23 (0.14–0.37) | <0.001 | 0.42 (0.23–0.75) | 0.004 |
| MACCE<sup>b</sup> | 0.56 (0.40–0.78) | <0.001 | 0.29 (0.19–0.44) | <0.001 | 0.52 (0.30–0.89) | 0.016 |
| Acute myocardial infarction | 0.34 (0.14–0.83) | 0.017 | 0.23 (0.09–0.61) | 0.003 | 0.67 (0.18–2.49) | 0.549 |
| Ischemic stroke | 0.78 (0.48–1.26) | 0.304 | 0.25 (0.12–0.54) | <0.001 | 0.33 (0.14–0.79) | 0.013 |
| Hemorrhagic stroke | 0.90 (0.49–1.64) | 0.721 | 0.86 (0.48–1.53) | 0.599 | 0.96 (0.42–2.17) | 0.914 |
| Heart failure hospitalization | 0.42 (0.27–0.66) | <0.001 | 0.19 (0.10–0.34) | <0.001 | 0.45 (0.21–0.93) | 0.031 |
| Advanced CKD | 0.52 (0.33–0.82) | 0.005 | 0.20 (0.11–0.39) | <0.001 | 0.39 (0.18–0.86) | 0.020 |
| ESRD with chronic dialysis | 0.67 (0.42–1.06) | 0.084 | 0.23 (0.11–0.46) | <0.001 | 0.34 (0.15–0.78) | 0.011 |
| Composite renal outcome | 0.58 (0.38–0.89) | 0.013 | 0.19 (0.10–0.37) | <0.001 | 0.33 (0.15–0.72) | 0.005 |

Abbreviation: CI, confidence interval; CKD, chronic kidney disease; ESRD, end-stage renal disease; HR, hazard ratio; MACCE, major adverse cardiac and cerebrovascular events; MACE, major adverse cardiovascular events; MRA, mineralocorticoid receptor antagonists; PA, primary aldosteronism.
a Any of cardiovascular death, acute myocardial infarction, and ischemic stroke.
b Any of cardiovascular death, acute myocardial infarction, ischemic stroke, and heart failure hospitalization.

